# Virtual Assessment of Patients with Dry Eye Disease During the COVID-19 Pandemic: One clinician’s experience

**DOI:** 10.1101/2022.03.07.22272050

**Authors:** Pierre Ibrahim, Caroline G. McKenna, Rookaya Mather

**Affiliations:** Western University, London, ON, Canada; BScH, Schulich School of Medicine and Dentistry, London, ON, Canada; MD, FRCSC, Schulich School of Medicine and Dentistry, Department of Ophthalmology, London, ON, Canada

**Keywords:** Dry Eye Disease, COVID, virtual care, quality of life, patient adherence

## Abstract

**Objectives:** To report on 1) the impact of DED on social, mental, and financial well-being, and 2) the use of virtual consultations to assess DED during the COVID-19 pandemic.

**Design & Methods:** An exploratory retrospective review of 35 charts. Telephone consultations for patients with DED conducted during the first lock-down period in Ontario in 2020 were reviewed.

**Results:** The most commonly reported DED symptoms were ocular dryness, visual disturbances, and burning sensation. The most common dry eye management practices were artificial tears, warm compresses, and omega-3 supplements. 20.0% of charts documented worsening of DED symptoms since the onset of the pandemic and 17.1% reported the lockdown had negatively affected their ability to perform DED management practices. 42.8% of patients reported an inability to enjoy their daily activities due to DED symptoms. 52.0% reported feeling either depressed, anxious, or both with 26.9% of patients accepting a referral to a social worker for counselling support. More than a quarter of the charts recorded financial challenges associated with the cost of therapy, and more than a fifth of patients reported that financial challenges were a direct barrier to accessing therapy.

**Conclusions:** Patients living with DED reported that their symptoms negatively affected their daily activities including mental health and financial challenges, that in turn impacted treatment practices. These challenges may have been exacerbated during the COVID-19 pandemic. Telephone consultations may be an effective modality to assess DED symptom severity, the impact of symptoms on daily functioning, and the need for counselling and support.

**AUTHOR SUMMARY:** Dry Eye Disease occurs when your tears do not provide enough lubrication for your eyes, which can be caused by either decreased tear production, or by poor quality tears. This study reviewed 35 patient charts to examine 1) the impact of Dry Eye Disease on patients’ well-being, and 2) the use of telephone appointments to assess Dry Eye Disease during the COVID-19 pandemic. Patients reported an inability to enjoy their daily activities due to symptoms of dry eye including burning sensation and blurred vision. Over half of patients reported mental health challenges. Over a quarter of patients reported that financial challenges prevented them from treating their Dry Eye Disease, such as affording eye drops, dietary supplements, and appointments to see their optometrist. These findings highlight that healthcare providers should considering quality of life, mental health, and financial challenges when treating patients with Dry Eye Disease. Through the experience of an ophthalmologist who specializes in Dry Eye Disease, telephone appointments may be an effective way to assess Dry Eye Disease symptoms, the impact of symptoms on daily functioning, and the need for counselling and support.

## INTRODUCTION

Dry eye disease (DED) is a multifactorial disease of the ocular surface characterized by loss of homeostasis of the tear film. Tear film instability and hyperosmolarity leads to ocular surface inflammation and damage(1), resulting in symptoms including burning and gritty sensation, photophobia, foreign body sensation, excess tearing, visual disturbances, and difficulty performing visual tasks(2). DED is a chronic and incurable disease, affecting an estimated six million Canadian adults(3). DED is known to negatively affect quality of life, work productivity(4), sleep(5), emotional well-being(2,6,7), and other everyday activities(8). Moreover, the negative impact of DED on daily living has been associated with mental health challenges such as depression and anxiety(9).

The COVID-19 pandemic has affected access to healthcare in Canada with devastating effects on healthcare systems globally. Canada’s pre-existing health care system was challenged with lengthy wait times(10) prior to the pandemic. Long wait times have been further exacerbated by the COVID-19 pandemic when lockdown periods resulted in cancelled or postponed appointments and delays in delivering care. In 2020, national wait times were longest for referral to ophthalmology compared to any other specialist referral with an average wait of 34.1 weeks(11).

Since the onset of the COVID-19 pandemic, many physicians have turned to virtual appointments as a solution in which physicians meet with patients via telemedicine modalities including phone and video chat. In Canada, virtual health care visits with primary care physicians and specialists jumped from 4% to 60% at the onset of the COVID-19 pandemic(10). There is existing literature presenting the use of virtual care for ophthalmic conditions such as glaucoma(12,13). During the Ontario COVID-19 lockdown period starting March 2020, non-urgent patient visits to the clinic were cancelled.

The objectives of this study are as follows: To report on 1) the impact of DED on patients’ social, mental, and financial well-being during the COVID-19 pandemic and 2) the experience of using virtual consultations to assess patients with DED during the COVID-19 pandemic.

## MATERIAL AND METHODS

### Study Design

Retrospective chart-review study from one cornea specialist practice at the Ivey Eye Institute, Schulich School of Medicine, Western University, London, Ontario, Canada. Patients had been referred by their optometrist or ophthalmologist for evaluation of dry eye disease. Only patients with adequate visual acuity (VA > 20/30) and normal intraocular pressure (IOP < 22mmHg), as documented by referring eyecare providers, were selected for virtual consultations.

Consultations were provided virtually over the phone using a structured questionnaire based on questions routinely asked in-person, and using validated questionnaires that are typically self-administered by patients during in-person consultations. The virtual consultation was divided into two parts; the first part was administered by a clinical assistant to obtain medical and ocular histories, document symptoms, current treatment, and CDEA (Canadian Dry Eye Assessment) and DEQ-5 (Dry Eye Questionnaire-5) scores in the same way that an ophthalmic technician would gather this information in a clinic setting. The second part of the DED consultation was conducted by telephone with the cornea specialist.

In light of the many disruptions to daily routines during the lockdown period, such as increased screen time from working from home, patients were asked whether their DED symptoms had changed during the lockdown and whether the lockdown was affecting their ability to engage in DED self-care. Clinical information was extracted from the charts and documented into data collection sheet with responses to 39 questions (S1 Appendix).

### Patient Population

All patients were referred by a physician or optometrist with an established diagnosis of DED. Patients were selected for virtual care appointments based on normal intraocular pressure and visual acuity, as reported by the referring physician or optometrist or ophthalmologist. A total of 35 patients had a virtual appointment booked. Six patients attended only the first part of the consultation. The data from first part of the consultation were included in the chart review.

### Data Analysis

Results are presented as a percentage of total responses collected for each question, as well as mean values of total responses. DED Management score (DEDM) is a composite score created to capture overall DED management practices using five indicators: use of artificial tears, use of tear gel, use of omega-3 supplements, use of warm compresses to the eyes, and use of eyelid cleansing. DEDM scores were tabulated by adding one point for each of the practices, leading to a scale from 0 to 5, where 5 indicates using all five management practices and 0 indicates using none the management practices. Spearman’s Rho tests were used to compare percentage values. JASP (v0.14.1) was used to conduct statistical analysis and results were considered statistically significant if P<0.05.

## RESULTS

### Participant Demographics

All charts (n= 35) recorded age, sex, general health history, including previously diagnosed Sjörgren’s syndrome, co-morbidities and systemic medications. All charts also documented ocular health history, dry eye history, ocular topical medications and other DED treatments, and frequency of optometric care. A total 35 patients completed the first part of the consultation interview with the clinical assistant. While not all patients answered every question, all completed questions were analyzed in the chart review as a proportion of those that did answer the question. On average, patients reported experiencing DED symptoms for 6.14 years prior to ophthalmology consultation and visiting their optometrist 1.61 times per year regarding DED (Table 1). A fifth of charts documented patients with a pre-existing diagnosis of Sjögren’s syndrome. 78.13% of charts documented a DEQ-5 score equal to or greater than 12, suggesting Sjögren’s related DED and warranting further work-up. 21.21% of charts documented severe DED symptoms and 48.48% had moderate DED symptoms according to CDEA scores equal to or greater than 31.

**Table 1:** Participant Demographics

| **Table 1: Participant Demographics** | **N** | **% or Mean** |
| --- | --- | --- |
| Total participants | 35 |  |
| Years living with DED symptoms (mean) |  | 6.14 years |
| Visits to doctor for DED complaints yearly | 35 |  |
| Mean |  | 1.61 +/-1.37 |
| More than once per year | 14 | 40.00% |
| Previous Sjögren's diagnosis | 35 |  |
| Yes | 7 | 20% |
| No | 28 | 80% |
| Dry Eye Questionnaire (DEQ-5) Score | 32 |  |
| Mean |  | 13.875 |
| Sjogren's (DEQ-5 > 12) | 25 | 78.13% |
| Canadian Dry Eye Assessment (CDEA) Score | 33 |  |
| Mean |  | 23.700 |
| Mild (CDEA 5-20) | 10 | 30.30% |
| Moderate (CDEA 21-30) | 16 | 48.48% |
| Severe (CDEA 31-48) | 7 | 21.21% |

### DED symptoms and management practices

The three most bothersome DED symptoms were documented for each patient. Results are shown in Fig 1. The worst symptoms experienced were ocular dryness (37.14%), blurred/fluctuating vision (20.00%), and burning/stinging (14.29%).

**Fig 1a.**
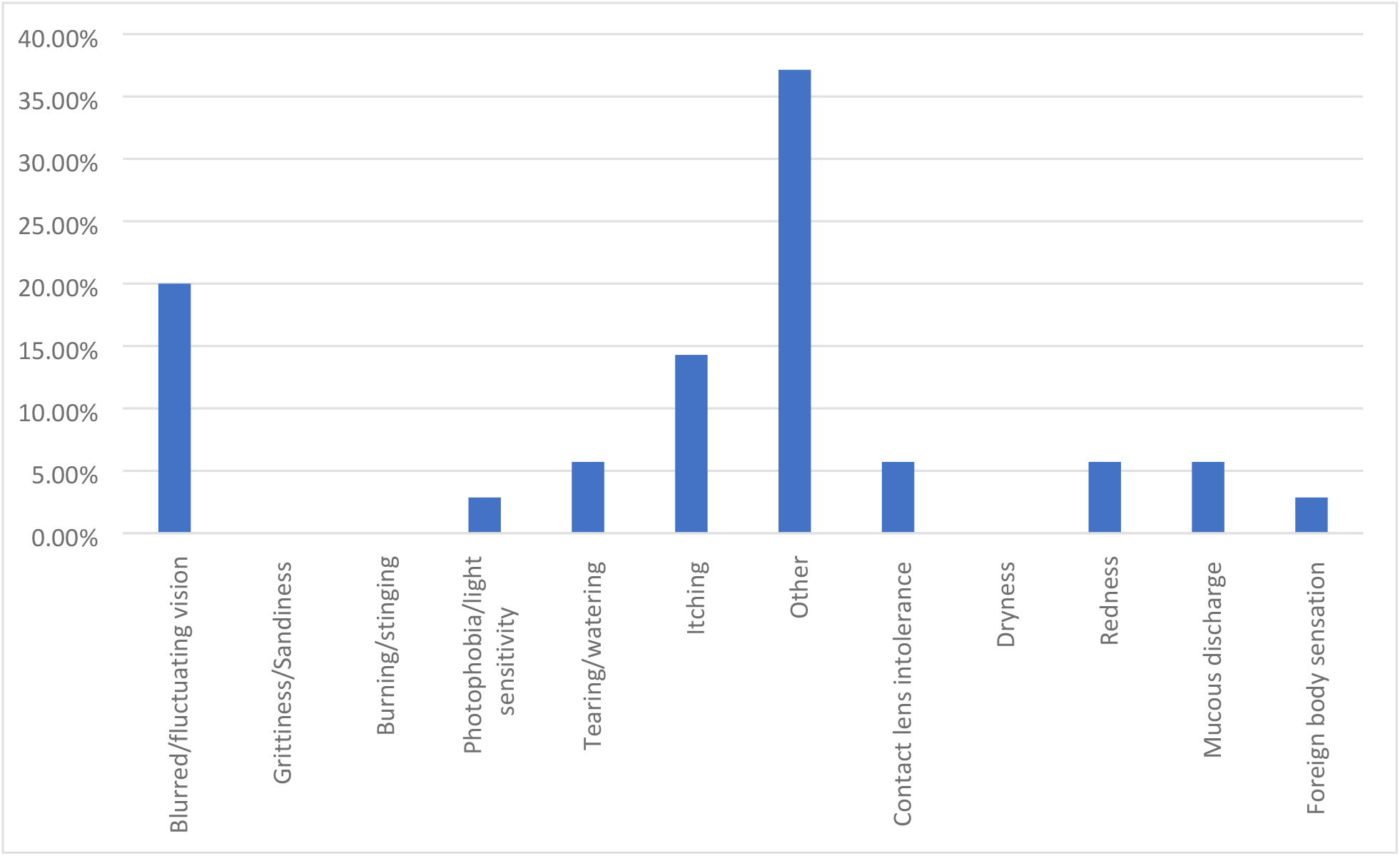
DED symptoms rated as most bothersome.

**Fig 1b.**
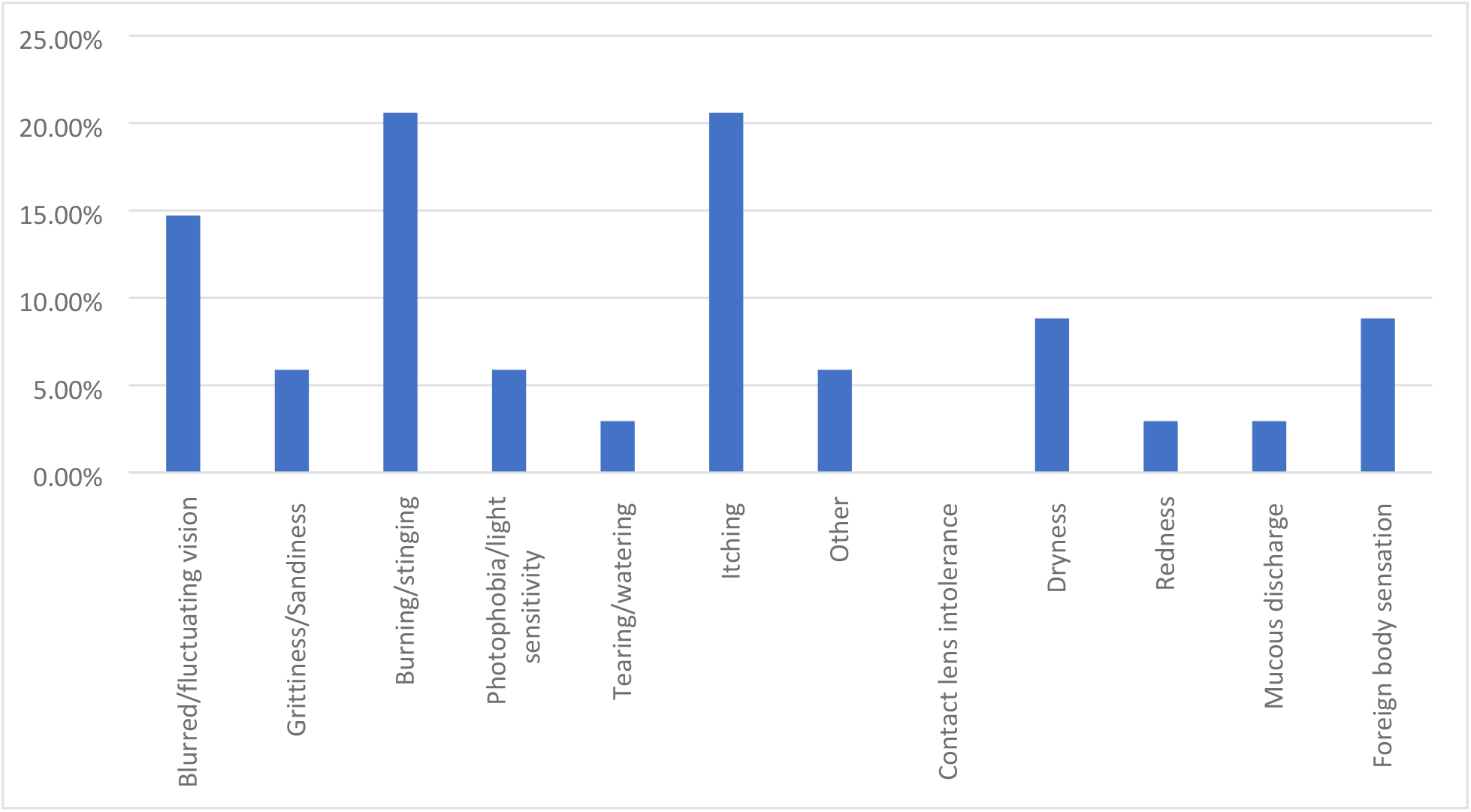
DED symptoms rated as second most bothersome.

**Fig 1c.**
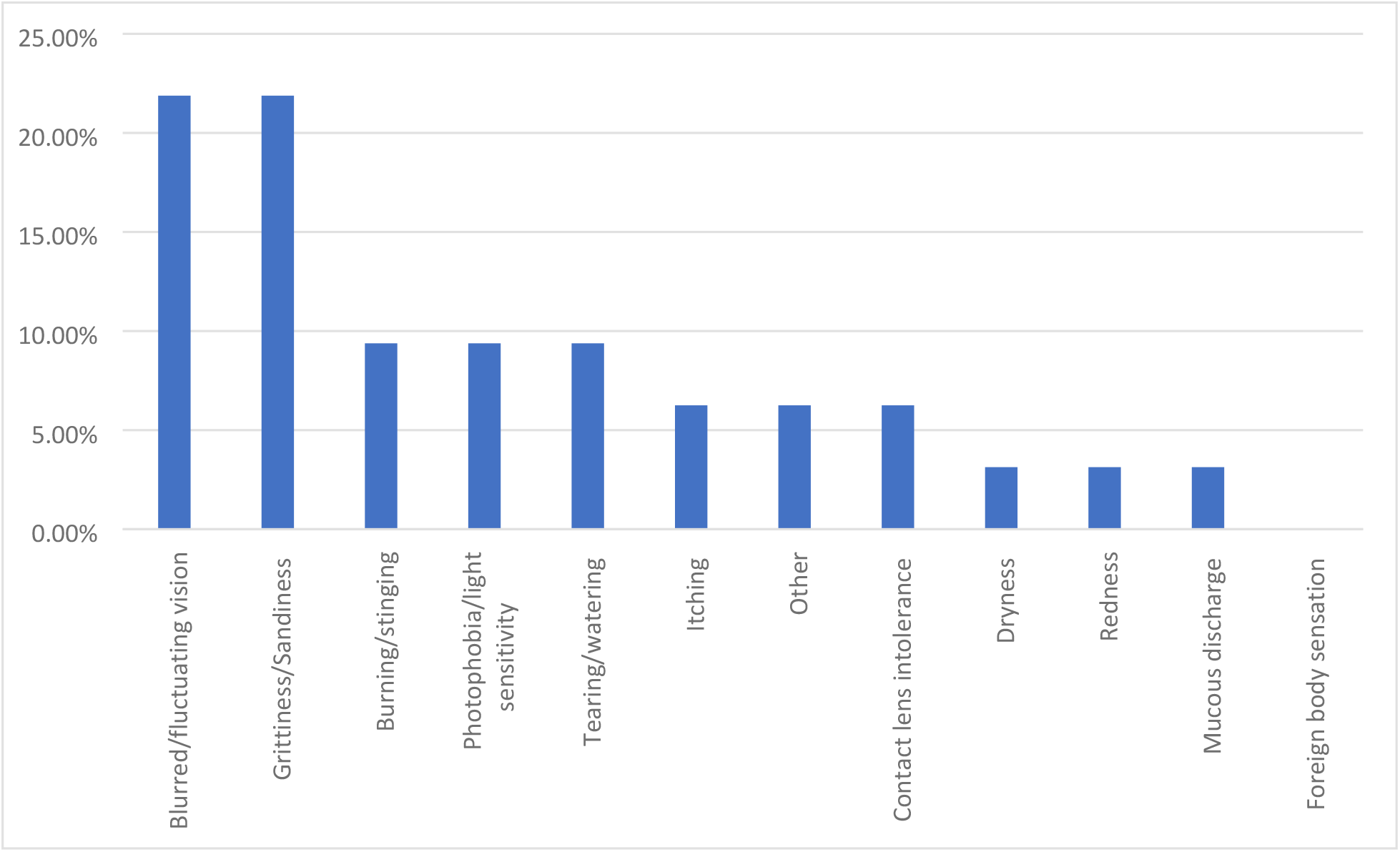
DED symptoms rated as third most bothersome.

The most common management practices among patients were artificial tear use (82.86%), warm compresses (71.43%), and omega-3 supplements (42.86%) (Table 2). However, among artificial tear users, 52.17% of charts documented the reported use to be less than three times per day. Among omega-3 users, 30.77% of charts documented taking less than the recommended daily dose of 2000 mg.

**Table 2:**
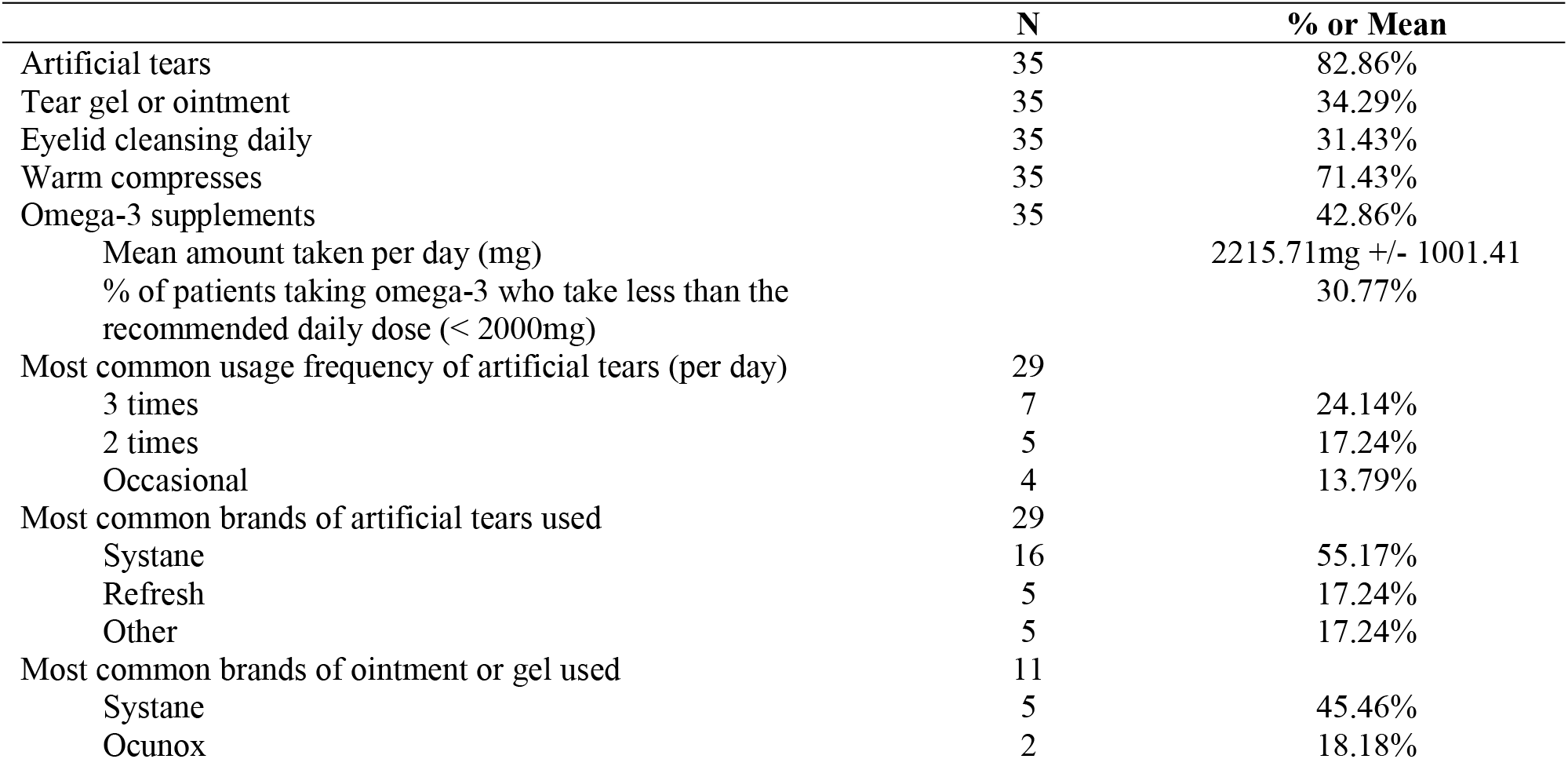
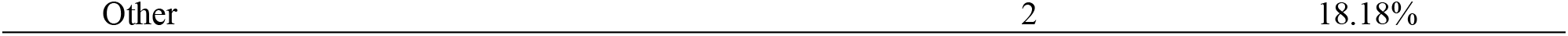
DED Management Practices

Spearman’s Rho analysis revealed that there was no significant correlation between DEDM scores and DEQ-5 scores (Table 3). However, there were significant correlations between number of artificial tears used per day and DEQ-5 score (P=0.009), between number of artificial tears per day and DEDM scores (P<0.001), and between number of artificial tears used per day and number of visits to eyecare provider per year (P=0.008).

**Table 3:** Spearman’s Rho Analysis

| Variables | CDEA Score | DEQ-5 Score | DEDM Score | Number Of Visits Per Year | Years with Symptoms | Number of Artificial Tears Per Day |
| --- | --- | --- | --- | --- | --- | --- |
| **CDEA Score** | - | - | - | - | - | - |
| **DEQ-5 Score** | Rho=0.527<br>$p=0.002^*$ | - | - | - | - | - |
| **DEDM Score** | Rho=0.192<br>$p=0.285$ | Rho=0.320<br>$p=0.070$ | - | - | - | - |
| **Number of Visits Per Year** | Rho= 0.176<br>$p=0.327$ | Rho= 0.122<br>$p= 0.499$ | Rho= 0.221<br>$p= 0.217$ | - | - | - |
| **Years with Symptoms** | Rho= 0.245<br>$p= 0.170$ | Rho= 0.350<br>$p= 0.046$ | Rho= 0.173<br>$p= 0.335$ | Rho= -0.076<br>$p= 0.673$ | - | - |
| **Number of Artificial Tears Per Day** | Rho= 0.183<br>$p= 0.309$ | Rho= 0.449<br>$p= 0.009^*$ | Rho= 0.704<br>$p= <0.001^*$ | Rho= 0.453<br>$p= 0.008^*$ | Rho= 0.110<br>$p= 0.542$ | - |

\* indicates $P<0.05$

### Impact of DED on daily activities, mental health, and finances

42.80% of charts documented patients reporting an inability to enjoy their daily activities due to DED symptoms (Table 4). The top three activities impacted by DED were reading (30.43%), spending time at the computer or watching TV (26.08%), and driving (17.39%) (Fig 2). As part of routine DED consultations, patients were screened for mental health challenges and asked to comment on the severity of their challenges as well whether they wished to be referred to social work team at the hospital for additional support. 48.00% of charts reviewed indicated no mental health difficulties. 52.00% of participants reported feeling either depressed, anxious, or both and 26.92% had accepted a referral to speak with a social worker when offered. More than a quarter of the charts documented patients reporting financial challenges associated with their DED. Similarly, more than a fifth of patients revealed that financial challenges were a barrier to accessing DED therapy.

**Table 4:**
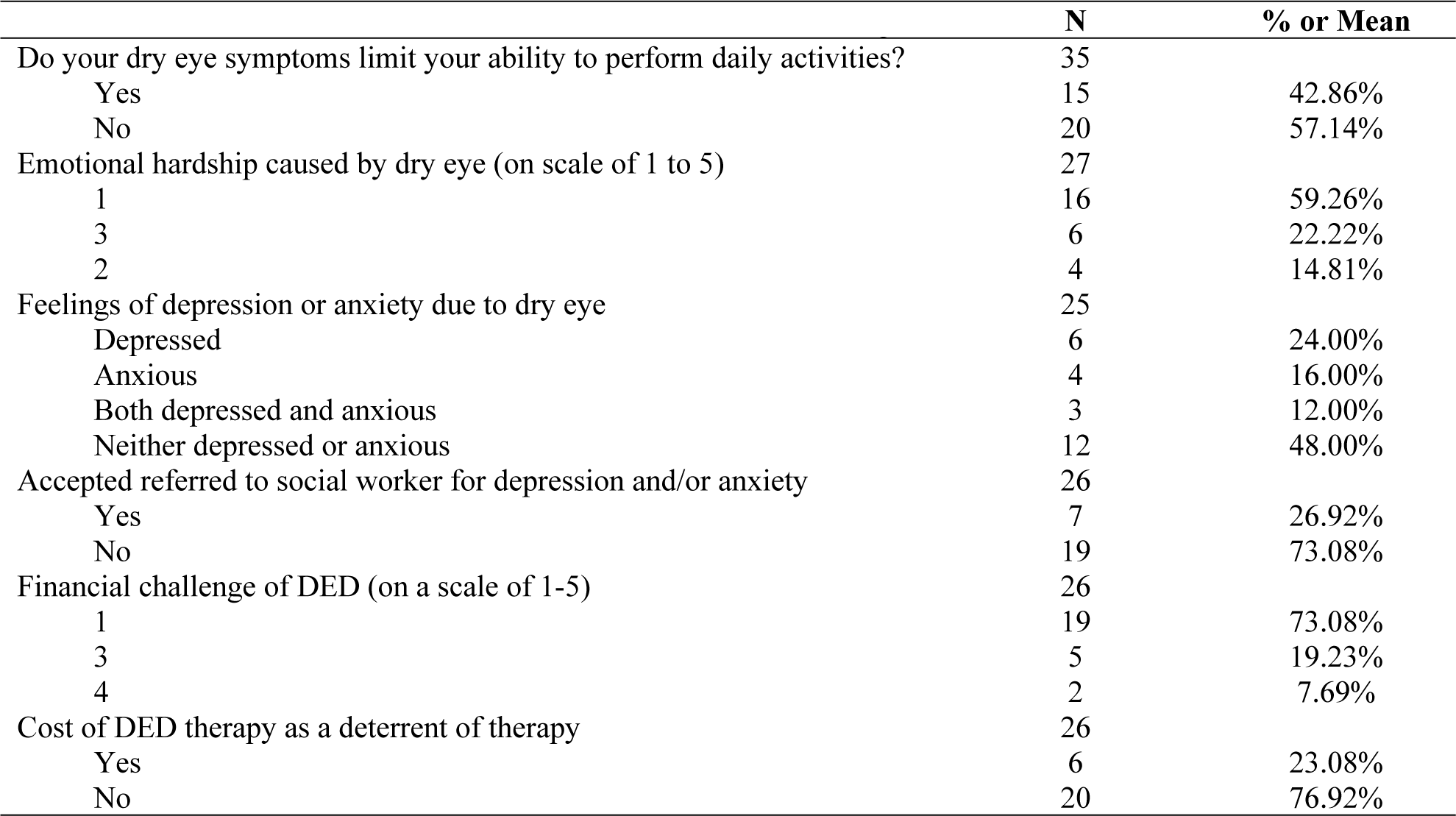
Effect of DED on mental, emotional, and financial well-being

| **Table 4: Effect of DED on mental, emotional, and financial well-being** | **N** | **% or Mean** |
| --- | --- | --- |
| Do your dry eye symptoms limit your ability to perform daily activities? | 35 |  |
| Yes | 15 | 42.86% |
| No | 20 | 57.14% |
| Emotional hardship caused by dry eye (on scale of 1 to 5) | 27 |  |
| 1 | 16 | 59.26% |
| 3 | 6 | 22.22% |
| 2 | 4 | 14.81% |
| Feelings of depression or anxiety due to dry eye | 25 |  |
| Depressed | 6 | 24.00% |
| Anxious | 4 | 16.00% |
| Both depressed and anxious | 3 | 12.00% |
| Neither depressed or anxious | 12 | 48.00% |
| Accepted referred to social worker for depression and/or anxiety | 26 |  |
| Yes | 7 | 26.92% |
| No | 19 | 73.08% |
| Financial challenge of DED (on a scale of 1-5) | 26 |  |
| 1 | 19 | 73.08% |
| 3 | 5 | 19.23% |
| 4 | 2 | 7.69% |
| Cost of DED therapy as a deterrent of therapy | 26 |  |
| Yes | 6 | 23.08% |
| No | 20 | 76.92% |

**Fig 2a.**
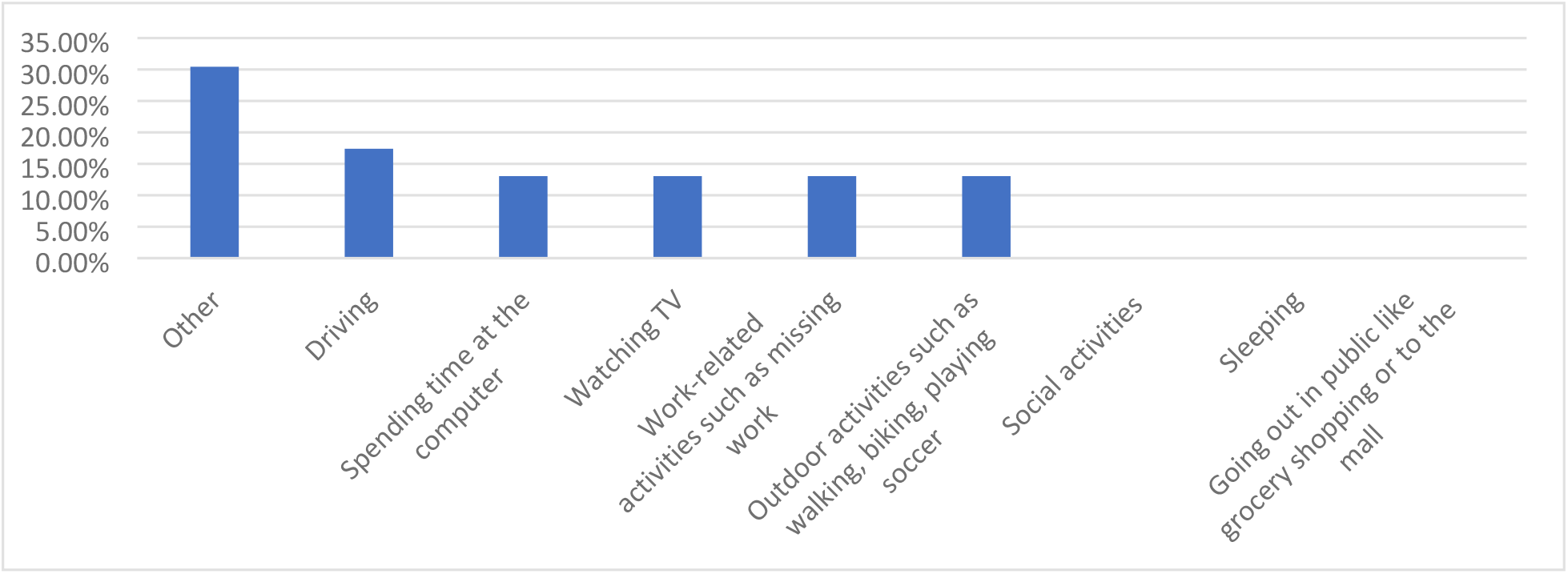
Most commonly affected activities due to DED.

**Fig 2b.**
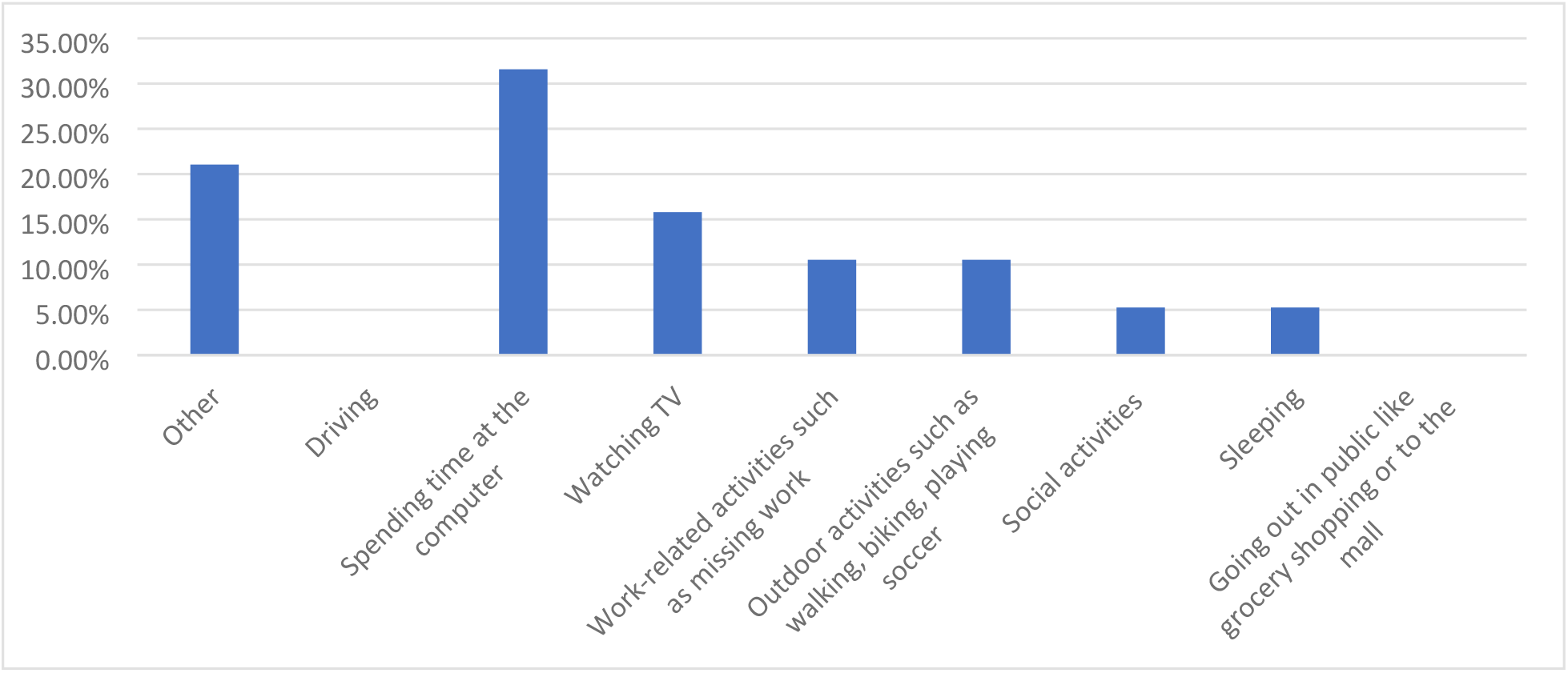
Second most affected activities due to DED symptoms.

**Fig 2c.**
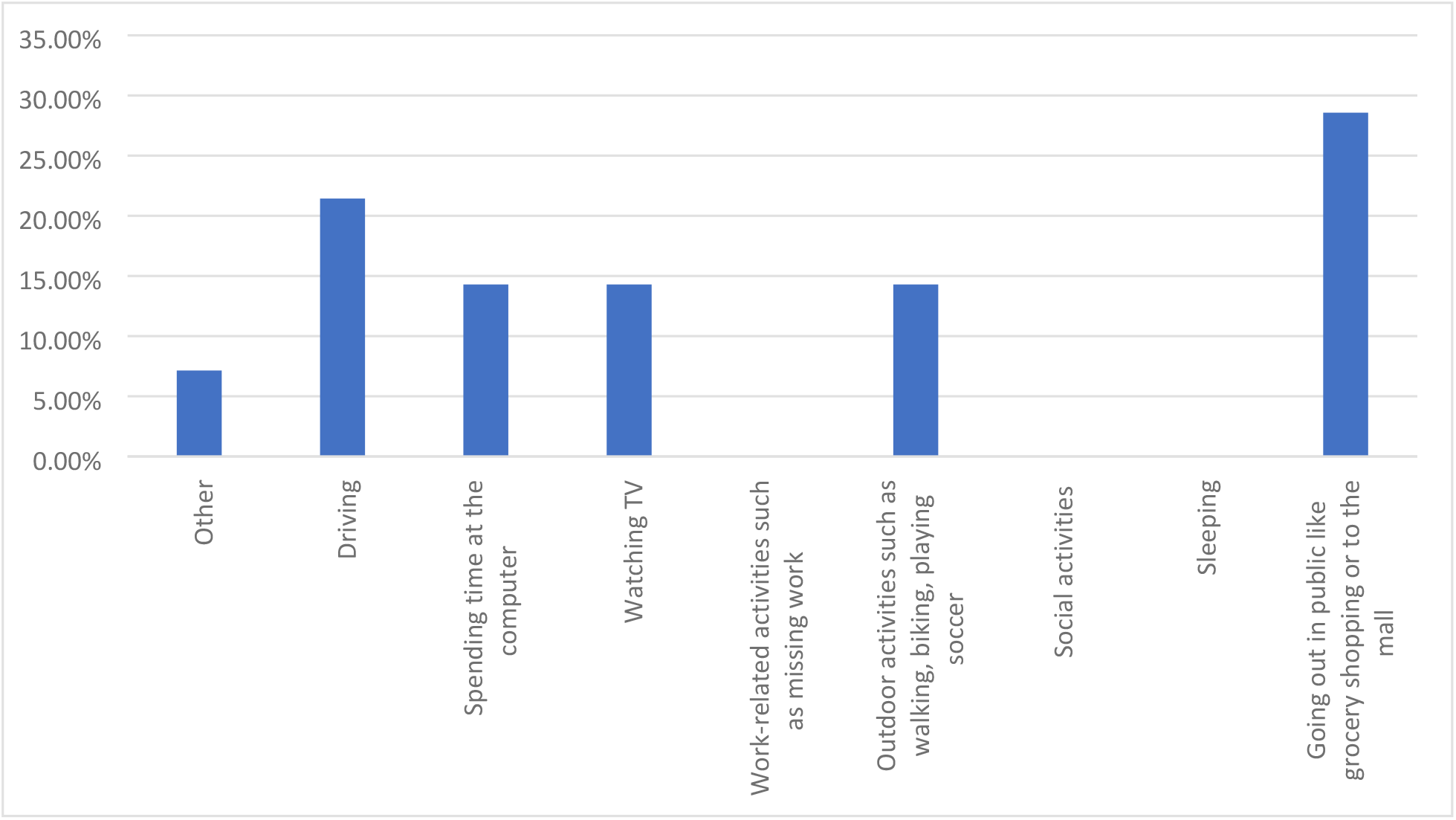
Third most affected activities due to DED symptoms.

### Impact of COVID-19 lockdown on DED

One fifth of charts documented worse DED symptoms during the lockdown period compared to prior to the lockdown period (Table 5). 17.14% of patients reported the lockdown negatively affected their ability to perform their DED self-care routine, 2.00% reported their self-care routine was better during the lockdown, and the remaining indicated no change.

**Table 5:**
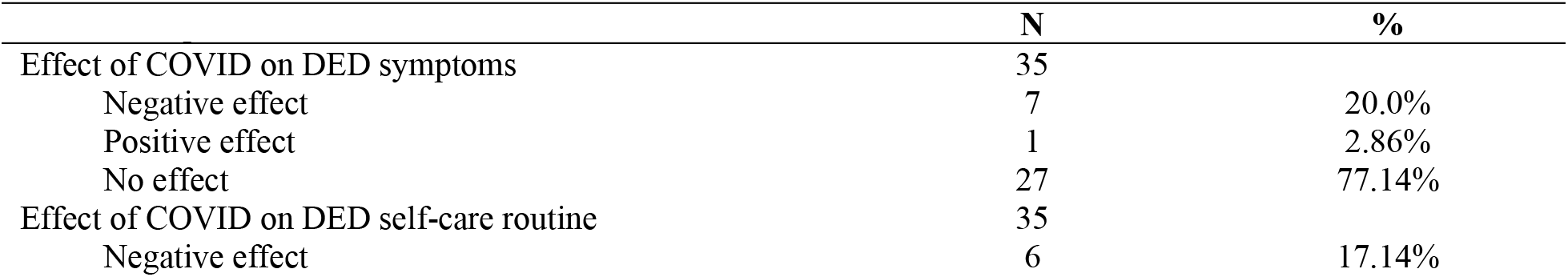
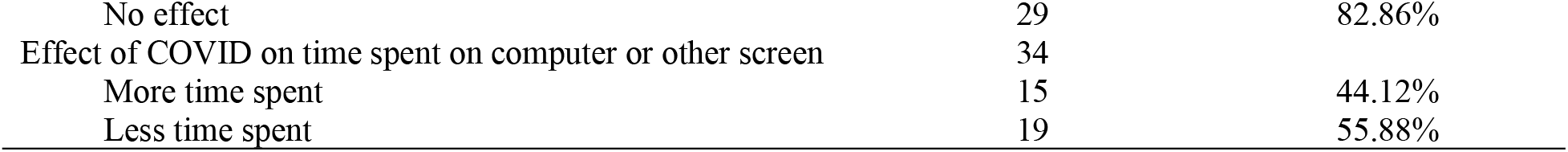
Impact COVID-19 on DED

## DISCUSSION

In this study, telephone consultations provided clinically useful information, allowing the cornea specialist to assess and grade patients’ current DED symptoms, impact on activities of daily living and mental health, and success in engaging in DED self-care activities. In addition, the virtual format allowed patients to disclose challenges requiring additional support to cope with DED. Virtual consultations identified patients warranting further investigations for Sjogren’s disease. In terms of treatment, telephone consultations allowed for initiation of optimal DED management, and allowed for triaging patients for subsequent in-person assessment.

### DED symptoms and self-care

In this preliminary chart review, patients were referred to a cornea specialist after an average of 6 years of DED symptoms, and almost three quarters of patients had moderate or severe DED symptoms according to CDEA scores. The majority of patients reported using artificial tears and warm compresses, and less than half reported using omega-3 supplements. Among those who used artificial tears, many were using drops less than four times per day despite the severity of symptoms reported. Among those who took omega-3 supplements, approximately one third were taking less than the 2000 mg recommended daily dose. With the information obtained, patients were counselled regarding optimal DED self-management and educated regarding optimal follow-up and future treatment modalities.

The DEDM score is a novel parameter created to capture overall DED management practices employed by patients. This measure allows researchers to understand how many of the five main practices were employed, to get a sense of self-management in totality. The composite score is a novel way to assess the relationships between patient treatment behaviours/practices and disease severity.

A significant correlation was identified between the frequency of artificial tear use and DEQ-5 scores. This was expected as worse symptoms intuitively warrant more tear use per day. In addition, the frequency of artificial tear use was correlated with overall DED management composite scores. Perhaps respondents who use tears more often are also more diligent about other elements of their DED self-care, or have the financial means to engage in more elements of self-care. The frequency of artificial tear use correlated with the number of eyecare visits per year, suggesting that more severe DED symptoms may lead to greater healthcare seeking behaviour. Financial means and/or insurance coverage allow patients to afford the cost of both artificial tears and additional eye care appointments. Moreover, seeing an optometrist more frequently may increase the likelihood of adherence to therapy.

### DED and financial barriers to therapy

Financial barriers associated with therapy may lead to poor adherence to DED self-care. Nearly a quarter of patients reported that the cost of dry eye therapy was a deterrent to therapy adherence. This is consistent with other studies that reported the cost of DED therapy represented an average annual out of pocket expense of $1,089 CAD per year(14) and $678-$1,267 USD per year(15). Moreover, this study found patients to be underutilizing artificial tears and omega-3 supplements. Financial barriers promote rationing of artificial tears and other therapeutics, as reported by Michaelov *et al*. (14). In addition, financial struggles are likely underreported by patients to eye care providers as previous studies have shown that non-adherence to therapy due to financial reasons is often not disclosed by patients to their health care providers(14). Furthermore, the COVID-19 pandemic has resulted in financial stressors for a large proportion of the population, as reported in a study that found 40% of US adults experienced COVID-19-related financial stressors from March to August 2020(16).

In this study population, patients visited their optometrist an average of 1.61 times per year. Interestingly, there was no correlation between frequency of visits and symptom severity. For some patients, insurance coverage may limit visits to once per year. The lack of private insurance coverage may also be a financial barrier to care. Literature demonstrates that lack of government-insured optometric services was found to negatively impact patients’ access to health care services, and ultimately vision health outcomes(17). However, these would be an important relationships to evaluate in a larger study sample.

### DED and mental health

In this study, over half of patients reported either anxiety, depression, or both, as a result of DED symptoms. This may be directly associated with symptoms of dry eye, or associated with difficulty performing daily activities due to DED symptoms. This is consistent with literature demonstrating the association between DED and mental health issues including anxiety and depression(6,18).

Over a quarter of patients accepted a referral to a social worker for support regarding living with DED. These findings highlight the importance of inquiring about mental health during DED appointments and may inform the rationale behind incorporating mental health screening into the standard of care for DED. This is essential given the high prevalence of mental health concerns among DED patients(4,7). Optometrists and ophthalmologists should not only prioritize inquiring about mental health, but also ensure they have the appropriate resources to provide to patients, such as a referral to a social worker, other mental health services, or to primary care providers.

Moreover, poor mental health may be exacerbated by the COVID-19 pandemic. There was a marked increase in the number of Canadians reporting ‘poor or fair’ mental health between March and May 2020, over half of which reported the decline in mental health was directly associated with lockdown measures that resulted in social isolation(19). In addition, the Canadian Mental Health Association reported Canadians were more likely to believe their mental health had worsened compared to physical health during the pandemic(20). While this study was not designed to evaluate an association or temporality between mental health, DED, and COVID-19, we acknowledge the possibility that there may be interactions between these variables.

### DED symptoms and the COVID-19 pandemic

One fifth of patients reported the pandemic made their DED symptoms worse, and 17% reported it negatively affected their ability to perform their DED self-care. Only 2% reported their symptoms improved. Nearly half of patients reported spending more time using screens, which may be contributing to the worsening symptoms.

Many non-essential medical visits and procedures were postponed due to provincial public health directives. In many cases, this resulted in a negative impact on health outcomes. Through virtual consultations, the patients in this study were assessed and counselled with respect to optimizing their DED self-care management. Despite the lack of physical exam findings such as VA, IOP, and SLE, virtual appointments may serve as a valuable way to initiate management until in-person visits are possible. Moreover, telephone appointments constitute a form of virtual care that doesn’t involve apps or video chat, making the modality user-friendly for both clinicians and patients. This achieves a similar goal as in person appointments when specialists provide counseling to patients and advice to referring providers.

Other forms of virtual care have been employed in ophthalmology. For example, in glaucoma virtual care, patients have been given handheld tonometers to measure IOP remotely to supplement video chat appointments(21). Others are using specialized video chat equipment to allow for improved visualization of the eye during virtual appointments. Recommendations for virtual care within the Canadian healthcare system include introducing education on telemedicine for medical students, and training in telemedicine for physicians with regards to delivery of virtual care, performing virtual patient examinations, and incorporating virtual care into their existing medical practices(22).

### Limitations

This study had a limited sample size due to the study objectives: to provide preliminary exploratory data to examine patient experience with DED during the COVID-19 pandemic, and report on the use of telephone consultations for DED referrals. Further studies are needed to better understand the effect of the COVID-19 pandemic on DED patients, as well as to evaluate the use of telephone, or other virtual care modalities, for DED consultations. Secondly, there are limitations to the composite scoring used in data analysis to capture how many elements of self-care patients were engaging in. The composite score weighed each self-care practice equally and without regard for variation, for example using one drop of artificial tears per day versus four times per day. In addition, not all therapies, or frequencies of therapies, are necessarily equally effective.

Finally, there are limitations to the virtual care modality itself. There were communication challenges over the telephone with some patients. This was especially evident in elderly patients, patients who were hard of hearing, and patients with language barriers. With regards to ophthalmological considerations, physical exam parameters such as VA, IOP, and SLE are impossible to conduct virtually. Though the most recently documented VA and IOP values were obtained from optometrist and ophthalmologist referral letters, the lack of contemporaneous VA, IOP, and SLEs may represent a limitation to care.

## CONCLUSIONS

Telephone consultations for patients with DED may be an effective way to assess dry eye disease symptom severity, the impact of symptoms on daily functioning, and the need for counselling and support. In this chart review, patients living with DED reported symptoms that negatively affected their daily activities including mental health and financial challenges, which in turn impact their ability to perform dry eye management activities. These challenges may have been exacerbated during the COVID-19 pandemic lockdown. Despite challenges and limitations, it is evident that simple virtual care via telephone will continue to play an integral role in DED care in the future.

## Data Availability

All relevant data are within the manuscript and its Supporting Information files.

## S1 APPENDIX. Virtual Dry Eye Consultation during the COVID-19 Pandemic

### History of the Presenting Complaint

1. What are the 3 most troubling eye symptoms you are experiencing? (List most troubling symptom, second most troubling symptom, third most troubling symptom from the list below)
  a. Dryness
  b. Grittiness/Sandiness
  c. Burning/stinging
  d. Foreign body sensation
  e. Photophobia/light sensitivity
  f. Contact lens intolerance
  g. Redness
  h. Mucous discharge
  i. Tearing/watering
  j. Itching
  k. Blurred/fluctuating vision
  l. Other
2. How long have you had dry eye symptoms (years)?
3. How many times per year do you visit your eye doctor regarding your dry eye?
4. Do you experience blurred vision along with symptoms of dryness?
5. Have you been diagnosed with a condition called Sjogren’s Syndrome?
6. Do you feel that your mouth is excessively dry?

### Past Ocular Therapy

7. Do you use eye drops for your dry eye condition?
8. How often do you use your drops per day?
9. What brand of drops do you use?
  a. Refresh
  b. Systane
  c. Liposic
  d. Hydrasense
  e. Hylo
  f. Soothe
  g. Other
10. Do you use tear gel or ointments?
11. How often do you use tear gel or ointments per day?
12. What brand of tear gel or ointment do you use?
  a. Refresh/Lacrilube
  b. Ocunox
  c. Systane
  d. Liposic
  e. Teargel
  f. Other
13. Do you perform eyelid cleansing daily?
14. Do you use warm compresses on your eyelids?
15. Do you take omega-3 supplements daily?
16. How much omega-3 do you take daily (mg)?
17. Have you had a chance to read the Dry Eye information package we mailed out to you?
18. DEQ-5 score (/22)?
19. CDEA score (/48)?
20. How many hours per day do you use a computer or other electronic device?
21. Do you use a C-PAP machine?

### Impact of DED Symptoms on Activities

22. Does your dry eye condition limit your ability to perform your daily activities?
23. List the top 3 activities you feel less able to enjoy or do because of your dry eye symptoms.
  a. Driving
  b. Spending time at the computer
  c. Work-related activities such as missing work
  d. Social activities
  e. Watching TV
  f. Going out in public like grocery shopping or to the mall
  g. Outdoor activities such as walking, biking, playing soccer
  h. Sleeping
  i. Other
24. Please rate how financial challenges MAY affect your ability to take care of your eye condition on a scale from 1 to 5 {1 = no issues at all; 3 = I find artificial tears and supplements expensive but I do buy what I need; 4 = I find treatment expensive and can only afford some of what I need 5 = I cannot afford the treatment all}
25. Does the cost of dry eye therapy prevent or limit how well you are able to care for your eyes?
26. Please tell me about the effect Dry Eye has on your mental and emotional well-being {1 = no effect and 5 = severe effect}
27. Do you feel depressed and/or anxious due to your dry eye symptoms?
28. Would you like to talk to someone about how you are feeling? I can refer you to our social worker who has helped many people who are struggling to cope with their dry eye condition?
29. Do you feel the COVID-19 pandemic and shutdown period has had an effect on your Dry Eye symptoms? Better? Worse? No change?
  a. If worse, why?
30. Did you spend more time using an electronic device?
31. Did the COVID pandemic/lockdown affect your ability to perform your regular Dry Eye self-care routine? If yes, how?
32. Has COVID pandemic affected how well you are coping with your dry eye condition? Better? Worse? No change?
  a. If worse, in what way?

### Past Ocular History

33. Do you have any other eye conditions such as glaucoma or macular degeneration?
34. Have you had any eye surgery in the past including laser vision correction?
35. What medical conditions do you have?
36. What prescription medications do you take?
37. Do you have any medication allergies?
38. Would you like us to email you additional information?
39. Do you smoke cigarettes?

## STATEMENTS

### Ethical Standards

The manuscript does not contain clinical studies or patient data.

### Author Contributions

Conceptualization: Rookaya Mather; Methodology: Rookaya Mather; Formal analysis and investigation: Caroline McKenna, Pierre Ibrahim,; Writing - original draft preparation: Caroline McKenna, Pierre Ibrahim; Writing - review and editing: Rookaya Mather, Caroline McKenna, Pierre Ibrahim; Funding acquisition: N/A; Resources: Rookaya Mather; Supervision: Rookaya Mather

